# Small palpebral fissure as a significant risk factor for glaucoma surgery failure

**DOI:** 10.64898/2026.05.27.26354208

**Authors:** Nanami Okuzumi, Sotaro Mori, Kana Katakami, Yuto Iwaki, Mari Sakamoto, Yuko Yamada-Nakanishi, Makoto Nakamura

**Affiliations:** Division of Ophthalmology, Department of Surgery, Kobe University Graduate School of Medicine, Address: 7-5-1 Kusunoki-cho, Chuo-ku, Kobe, Hyogo, 650-0017, Japan

**Keywords:** glaucoma surgery, risk factor, small palpebral fissure, MIGS, filtering surgery

## Abstract

**Purpose:** To evaluate the impact of “not commonly considered risk factors” on glaucoma surgical outcomes.

**Methods:** This study included 339 eyes that underwent glaucoma surgery. Surgical procedures included microhook *ab-interno* trabeculotomy (µTLO), Preserflo *ab-externo* microshunt implantation, trabeculectomy (Trab), and Ahmed Glaucoma Valve (AGV) implantation. In addition to conventional background factors, we examined a set of “not commonly considered risk factors,” including very elderly age (≥85 years), avitreous status, aphakia, use of antithrombotic agents, difficulty attending frequent postoperative visits, small palpebral fissure, corneal endothelial dysfunction, poor vision in the fellow eye, dementia, hearing loss, mental illness, atopic dermatitis, pseudophacodonesis, glaucoma eye drop allergy, and conditions contraindicating β-blocker use. Surgical success was defined as intraocular pressure (IOP) ≤21 mmHg, ≥20% reduction from baseline, and no additional glaucoma surgery at 1 year. Logistic regression was performed to identify potential risk factors; significant factors were further evaluated using propensity score matching.

**Results:** Of the 339 cases, surgical success rates were 65% for µTLO, 82% for Preserflo, 91% for Trab, and 82% for AGV. Multivariate logistic regression identified two independent predictors of surgical failure: small palpebral fissure (odds ratio 2.52, p < 0.01) and hearing loss (odds ratio 3.94, p = 0.04). Propensity score matching of patients with small versus large palpebral fissures (111 per group) confirmed significantly worse postoperative outcomes in the small-palpebral-fissure group despite balanced baseline characteristics.

**Conclusion:** Small palpebral fissure is an independent and previously unnoticed risk factor for glaucoma surgical failure, affecting both minimally invasive and filtration procedures.

**Précis:** This study identifies small palpebral fissures as an independent risk factor for glaucoma surgery
failure and highlights the clinical importance of evaluating under-recognised patient characteristics
when planning surgical procedures.

## Introduction

In recent years, as society has matured and the global population has aged, the number of patients presenting with multiple comorbidities has increased.^1^ Advances in medicine have enabled more aggressive treatments for patients who previously had limited options due to severe systemic disease. However, these advances have also created a situation in which surgeons are required to manage cases with increasingly complex and heterogeneous background factors. In the field of glaucoma, in particular, the disease is chronic and progressive and occurs more commonly in elderly patients,^2^ making a multifaceted perspective essential when determining the appropriateness of surgery.

Factors known to influence the selection of surgical procedures and postoperative outcomes in glaucoma surgery have generally been described in large-scale studies.^3-5^ These include age, sex, race, preoperative intraocular pressure (IOP), number of preoperative glaucoma medications, glaucoma disease type, lens status, history of intraocular surgery, and concomitant cataract surgery. Such factors are typically treated quantitatively as “preoperative background factors” and form the basis of current evidence.

However, in clinical practice, many “not commonly considered risk factors” that are not captured by these standard parameters have substantial influence on the choice of surgical procedure and on the postoperative course. For example, procedures requiring postoperative management tend to be avoided in patients with dementia or mental illness. Long procedures are often avoided in patients with significant hearing loss or in the very elderly due to the risk of intraoperative agitation. In cases where the fellow eye has poor visual function, procedures that may temporarily reduce vision are approached with caution. Moreover, in patients with small palpebral fissures, conjunctival manipulation is difficult. As a result, filtering surgery becomes technically disadvantageous, and alternative approaches are often selected. Additionally, for patients who live far from the hospital and have difficulty attending postoperative visits, procedures requiring less intensive postoperative care may be preferred.

Thus, surgeons incorporate both evidence-based factors and non-quantitative considerations, including physical, social, and psychological aspects, when selecting the optimal surgical approach. Despite this, there has been little systematic evaluation of the extent to which these “not commonly considered risk factors” influence surgical outcomes. In an era where patient backgrounds are becoming increasingly diverse, overlooking these factors may create a significant gap in the accuracy of surgical decision-making and the interpretation of treatment results. Therefore, in this study, we focused on these “not commonly considered risk factors” and examined their impact on the outcomes of glaucoma surgery.

## Method

This study included glaucoma surgeries performed at Kobe University Hospital between October 2023 and October 2024, encompassing microhook *ab-interno* trabeculotomy (µTLO) Preserflo ab-externo microshunt implantation, trabeculectomy (Trab), and long tube shunt procedures. In bilateral cases, only the first operated eye was included. For patients who underwent multiple surgeries during the study period, only the initial glaucoma surgery was analysed. Cases lacking follow-up data through 1 year postoperatively were excluded. At our institution, only microhook *ab interno* trabeculotomy^6^ was performed as minimally/micro invasive glaucoma surgery (MIGS), and all long tube shunt surgeries used the Ahmed Glaucoma Valve (AGV) during this period. Details of each surgical technique have been described previously.^7-12^ The study adhered to the tenets of the Declaration of Helsinki and was approved by the institutional review board of Kobe University Hospital (No. B250117).

General patient parameters included age, sex, glaucoma disease type, glaucoma drug score (GDS), axial length, and history of intraocular surgery. The GDS was calculated by assigning 1 point per topical medication and 2 points for combination formulations or oral carbonic anhydrase inhibitors.^6, 9-12^ Surgical variables included surgeon and concomitant cataract surgery. Postoperative outcomes included IOP and GDS at 1 year after surgery. Surgical success was defined according to previous reports^6, 9-12^ as meeting all of the following criteria: (1) IOP ≤ 21 mmHg, (2) ≥20% reduction from preoperative IOP, and (3) no additional glaucoma surgery. Bleb revision surgery was not considered additional glaucoma surgery.

In this study, “not commonly considered risk factors” were defined as the presence of any of the following: very elderly age (≥85 years), ^13^ avitreous status, aphakia, use of antithrombotic agents, difficulty attending frequent follow-up visits, small palpebral fissure, corneal endothelial dysfunction with a preoperative endothelial cell density ≤1000/mm^2^,^14^ poor vision in the fellow eye, dementia, hearing loss, mental illness, atopic dermatitis, pseudophacodonesis, allergy to glaucoma medications, or asthma/heart disease precluding the use of β-blocker eye drops. Difficulty attending frequent visits was defined as (1) residing in an area requiring more than 2 hours of one-way door-to-door travel to our hospital, or (2) situations in which the patient or their family reported that frequent postoperative visits would be difficult due to social circumstances. A small palpebral fissure was defined as a conjunctival exposure width of ≤5 mm—measured as the shortest distance from the superior nasal limbus to the eyelid margin with the patient gazing outward and downward—prior to surgery (**Figure 1**). Poor vision in the fellow eye was defined as corrected logMAR visual acuity ≥0.4 in the non-operated eye.^15^ Dementia and hearing loss were included when these conditions were formally diagnosed and judged by the surgeon to potentially impair communication during surgery under local anaesthesia or during postoperative care. The prevalence of these factors for each surgical procedure and their association with surgical risk were assessed using logistic regression.

**Figure 1.**
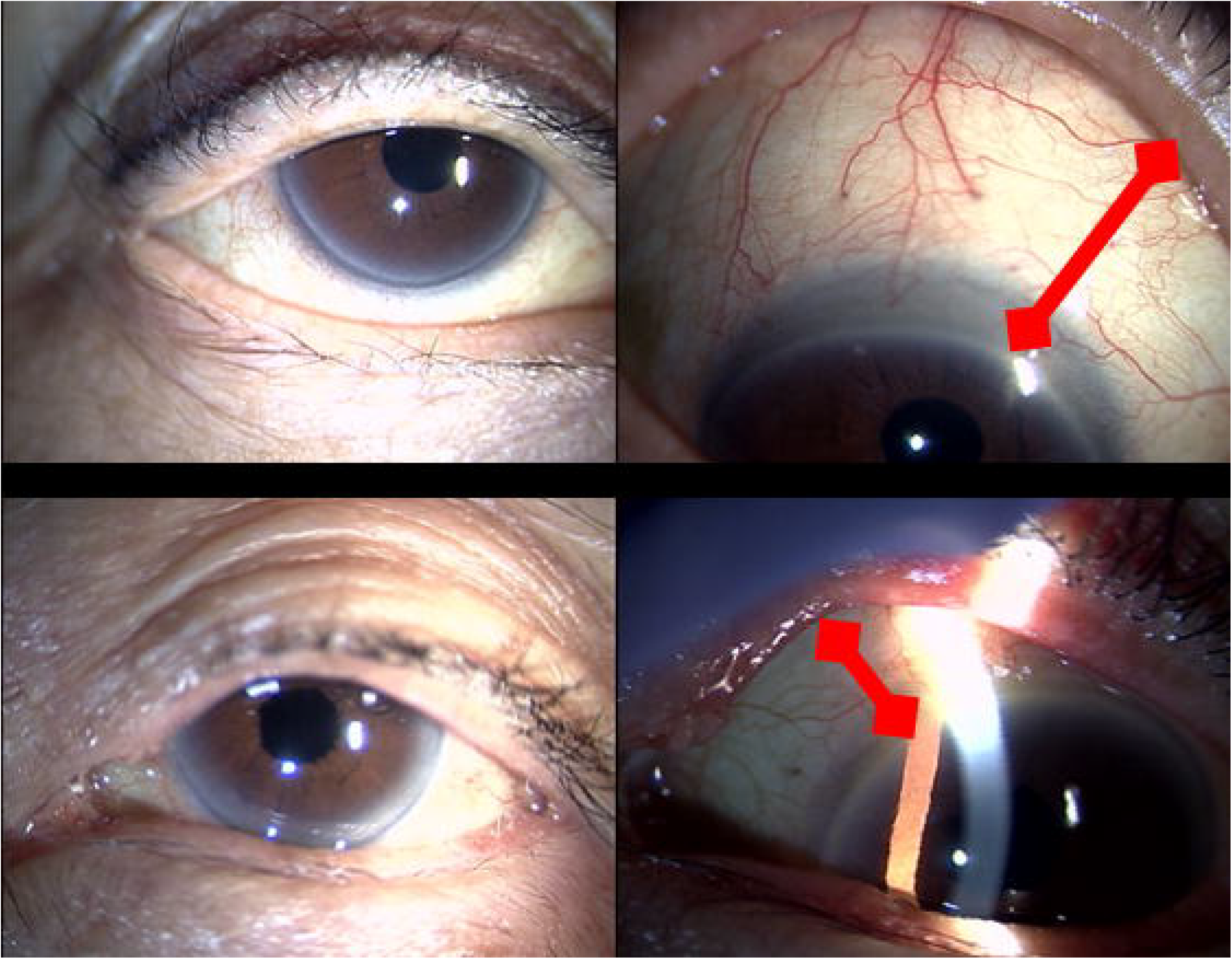
Measurement of the Palpebral Fissure. The distance from the corneal limbus to the nearest eyelid margin (arrow) was measured in a downward gaze. A small palpebral fissure was defined as ≤5 mm.

Logistic regression analysis was first performed to identify potential risk factors, including those not commonly evaluated in prior studies. As a sensitivity analysis, we conducted propensity score– matched analyses to assess whether these factors represented true risk factors. Propensity scores were estimated using logistic regression to calculate the probability of having each factor, with covariates including age, sex, glaucoma disease type, surgical procedure, surgeon, preoperative IOP, number of prior intraocular surgeries, and combined cataract surgery. A caliper width of 0.2 was applied, and 1:1 matching was performed.

## Results

**Table 1** summarises patient characteristics and postoperative outcomes. A total of 339 cases were included: 82 µTLO, 179 Preserflo, 56 Trab, and 22 AGV cases. The proportion of patients undergoing concurrent cataract surgery was highest in the µTLO group (61%), followed by the Preserflo group (32%). Compared with other surgical procedures, AGV cases demonstrated a higher proportion of other secondary glaucoma (55%) and a greater history of prior intraocular surgeries (median: 2.5). The overall surgical success rate was slightly lower for µTLO (65%), whereas filtering surgeries showed success rates exceeding 80%, with Trab achieving the highest rate (91%).

**Table 1.**
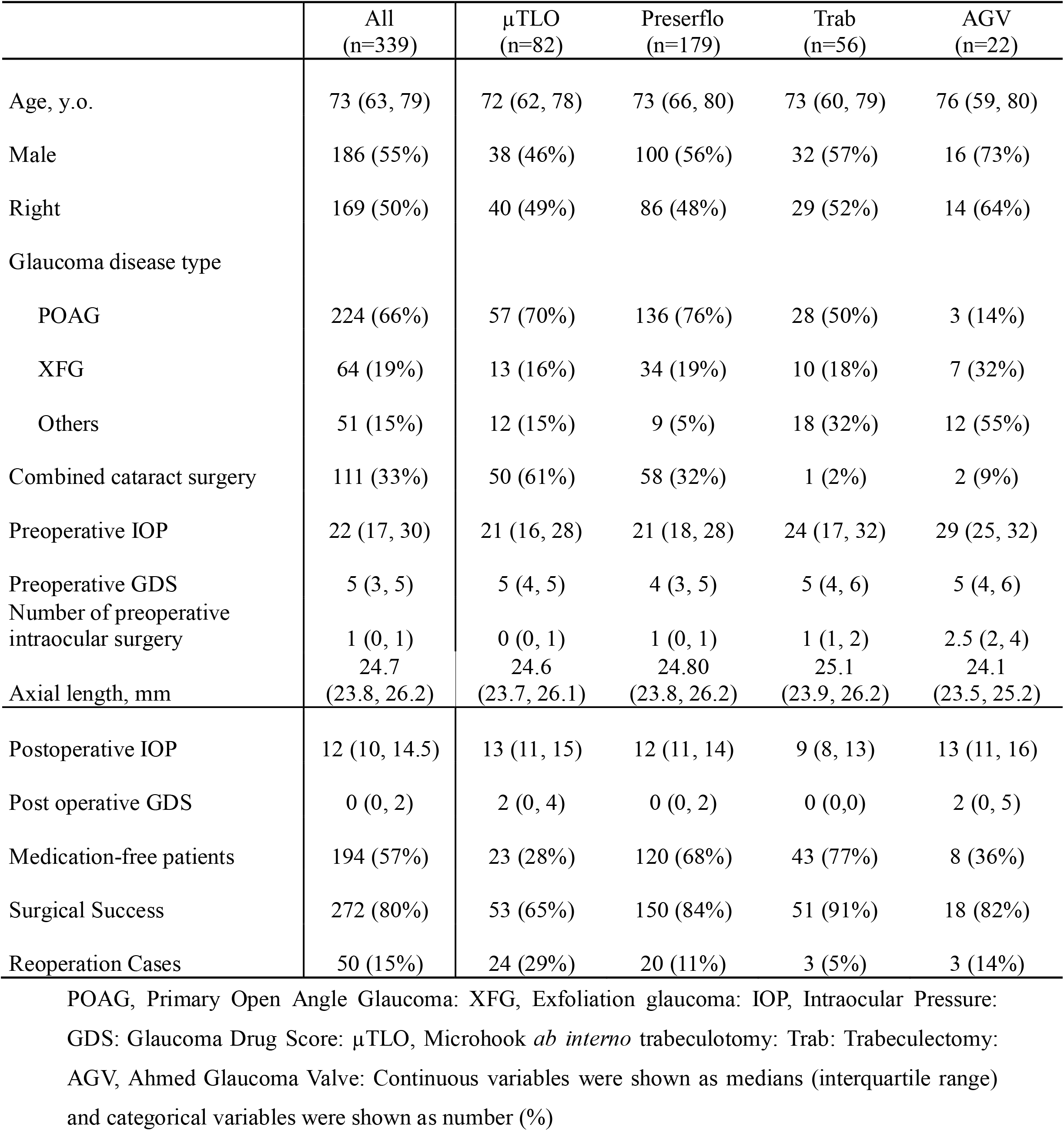
Patient characteristics and postoperative outcomes by surgical procedure.

**Table 2** presents the preoperative prevalence of “not commonly considered risk factors.” The most frequent factors were small palpebral fissure (38%), glaucoma medication allergy (32%), and difficulty with frequent visits (22%). The use of antithrombotic agents was more common in Preserflo than in Trab cases (23% vs. 5%). AGV surgery was associated with the highest overall prevalence of these risk factors compared with the other procedures. Among the four procedure groups, AGV also showed the highest rates of very elderly age (≥85 years), avitreous status, aphakia, antithrombotic agent use, difficulty with frequent visits, corneal endothelial dysfunction, poor vision in the fellow eye, atopic dermatitis, and asthma or heart disease.

**Table 2.** Prevalence of factors not commonly considered risk factors stratified by surgical procedure.

|  | All<br>(n=339) | µTLO<br>(n=82) | Preserflo<br>(n=179) | Trab<br>(n=56) | AGV<br>(n=22) |
| --- | --- | --- | --- | --- | --- |
| Very elderly age (≥85 Years) | 19 (6%) | 2 (2%) | 11 (6%) | 4 (7%) | **2 (9%)** |
| Avitreous | 18 (5%) | 1 (1%) | 4 (2%) | 1 (2%) | **12 (55%)** |
| Aphakia | 2 (1%) | 0 (0%) | 0 (0%) | 0 (0%) | **2 (9%)** |
| Antithrombotic agents | 59 (17%) | 7 (9%) | 41 (23%) | 3 (5%) | **8 (36%)** |
| Difficulty with frequent visits | 74 (22%) | 11 (13%) | 47 (26%) | 10 (18%) | **6 (27%)** |
| Small palpebral fissure | 128 (38%) | 31 (38%) | **72 (40%)** | 18 (32%) | 7 (32%) |
| Corneal endothelial disorder | 12 (4%) | 1 (1%) | 4 (2%) | 2 (4%) | **5 (23%)** |
| Poor vision in fellow eye | 67 (20%) | 11 (13%) | 39 (22%) | 10 (18%) | **7 (32%)** |
| Dementia | 5 (2%) | 0 (0%) | 4 (1%) | **1 (2%)** | 0 (0%) |
| Hearing loss | 12 (4%) | 3 (4%) | **8 (5%)** | 1 (2%) | 0 (0%) |
| Mental illness | 19 (6%) | **6 (7%)** | 10 (6%) | 3 (5%) | 0 (0%) |
| Atopic dermatitis | 22 (7%) | 6 (7%) | 11 (6%) | 2 (4%) | **3 (14%)** |
| Pseudophacodonesis | 4 (1%) | 1 (1%) | 1 (1%) | **2 (4%)** | 0 (0%) |
| Glaucoma eye drop allergy | 107 (32%) | 27 (33%) | **59 (33%)** | 17 (30%) | 4 (18%) |
| Asthma / Heart Disease | 50 (15%) | 10 (12%) | 28 (16%) | 6 (11%) | **6 (27%)** |

µTLO, Microhook *ab interno* trabeculotomy; Trab: Trabeculectomy; AGV, Ahmed Glaucoma Valve.
Bold indicates the procedure with the highest rate.

**Supplemental Table 1** shows the surgical success rates stratified by these risk factors. Among them, aphakia (50% failure; 1 of 2 cases), atopic dermatitis (64% failure; 14 of 22 cases), and hearing loss (67% failure; 4 of 12 cases) were associated with the highest rates of surgical failure. Furthermore, for most of these risk factors, success rates for µTLO were lower than the procedure-specific rates shown in Table 1.

**Table 3** shows the results of logistic regression analyses evaluating whether “not commonly considered risk factors” were associated with surgical success. Because 67 surgical failures occurred, inclusion of all risk factors in a multivariate model was not feasible. Therefore, univariate analyses were first performed, excluding corneal endothelial dysfunction, dementia, and mental illness, which were not expected to be strongly related to surgical success. Multivariate analysis was then conducted for the remaining factors, excluding aphakia and pseudophacodonesis due to their very low prevalence and excluding antithrombotic agent use because its impact had previously been examined in detail at our facility.^8^ In the multivariate analysis, small palpebral fissure (odds ratio 2.52, p < 0.01) and hearing loss (odds ratio 3.94, p = 0.04) emerged as significant risk factors for surgical failure.

**Table 3.** Logistic regression analysis of surgical outcomes for factors not commonly considered glaucoma risk factors.

|  | Univariate analysis |  |  | Multivariate analysis |  |  |
| --- | --- | --- | --- | --- | --- | --- |
|  | Odds ratio | 95%CI | P value | Odds ratio | 95%CI | P value |
| Very elderly age ($\geq 85$ Years) | 0.72 | 0.20-2.55 | 0.61 | 0.6 | 0.15-2.33 | 0.46 |
| Avitreous | 0.47 | 0.11-2.11 | 0.33 |  |  |  |
| Aphakia | 3.96 | 0.24-64.10 | 0.33 |  |  |  |
| Antithrombotic agents | 0.56 | 0.25-1.25 | 0.16 |  |  |  |
| Difficulty with frequent visits | 1.1 | 0.59-2.07 | 0.76 | 1.13 | 0.58-2.22 | 0.72 |
| Small palpebral fissure | 2.27 | 1.33-3.89 | **<0.01** | 2.52 | 1.44-4.40 | **<0.01** |
| Corneal endothelial disorder | 1.32 | 0.35-5.00 | 0.69 |  |  |  |
| Poor vision in fellow eye | 0.63 | 0.31-1.32 | 0.22 | 0.52 | 0.24-1.13 | 0.98 |
| Dementia | 0.97 | 0.11-8.86 | 0.98 |  |  |  |
| Hearing loss | 2.94 | 0.90-9.55 | 0.07 | 3.94 | 1.09 – 14.20 | **0.04** |
| Mental illness | 1.05 | 0.34-3.26 | 0.94 |  |  |  |
| Atopic dermatitis | 2.4 | 0.96-5.97 | 0.06 | 2.43 | 0.93-6.27 | 0.07 |
| Pseudophacodonesis | 0.25 | 0.14 – 1.49 | 0.98 |  |  |  |
| Glaucoma eye drop allergy | 1.2 | 0.69-2.10 | 0.52 | 1.29 | 0.72-2.30 | 0.39 |
| Asthma / Heart Disease | 1.12 | 0.54-2.33 | 0.75 |  |  |  |

Bold indicates p-values less than 0.05.

Given that small palpebral fissure was a significant risk factor in both univariate and multivariate analyses, we conducted a sensitivity analysis using propensity score matching to compare outcomes between patients with small versus large palpebral fissures (**Table 4**). Using 1:1 caliper matching, 111 patients were selected for each group. Although preoperative characteristics did not differ between the groups, the small palpebral fissure group had significantly worse postoperative outcomes across all metrics, including IOP, GDS, surgical success rate, and need for reoperation. There were also no significant differences between the two matched groups in the prevalence of other “not commonly considered risk factors” (**Supplemental Table 2**).

**Table 4.** Pre- and postoperative outcomes in the small vs. large palpebral fissure groups using propensity score matching.

|  | Small palpebral fissure<br>(n=111) | Large palpebral fissure<br>(n=111) | P value |
| --- | --- | --- | --- |
| Age, y.o. | 75(66, 81) | 73(63, 79) | 0.47 |
| Male | 58 | 64 | 0.50 |
| Right | 56 | 54 | 0.89 |
| Glaucoma disease type<br>(POAG, XFG, Others) | 77, 20, 14 | 72, 19, 20 | 0.55 |
| Surgery type<br>(μTLO, PMS, Trab, AGV) | 27, 63, 16, 5 | 29, 56, 20, 6 | 0.81 |
| Combined cataract surgery | 35 (32%) | 32 (29%) | 0.77 |
| Surgeon (A, B, C, D) | 27:38:18:28 | 29:40:20:22 | 0.82 |
| Preoperative IOP | 22 (18, 30) | 22(17, 28) | 0.91 |
| Preoperative GDS | 5 (4,5) | 4(3,5) | 0.23 |
| Number of preoperative intraocular<br>surgery | 1 (0,1) | 1(0,1) | 0.74 |
| Postoperative IOP | 12(10, 16) | 12(10, 14) | **0.03** |
| Post operative GDS | 1(0,3) | 0(0,2) | **<0.01** |
| Medication-free patients | 55 (50%) | 73 (66%) | **0.02** |
| Surgical success | 79 (71%) | 94 (85%) | **0.02** |
| Reoperation cases | 26 (23%) | 11 (10%) | **0.02** |

POAG, Primary Open Angle Glaucoma: XFG, Exfoliation glaucoma: μTLO, Microhook *ab interno* trabeculotomy: Trab: Trabeculectomy: AGV, Ahmed Glaucoma Valve: IOP, Intraocular Pressure: GDS: Glaucoma Drug Score: Bold indicates p-values less than 0.05. Continuous variables were shown as medians (interquartile range) and categorical variables were shown as number (%).

## Discussion

This study identified small palpebral fissures and hearing loss as contributors to glaucoma surgery failure among the “not commonly considered risk factors” that have not been sufficiently evaluated in previous research. Small palpebral fissures, in particular, consistently emerged as a significant risk factor in both logistic regression and propensity score–matched analyses, highlighting the strength of their impact. Previous reports have shown that severe prostaglandin-associated periorbitopathy (PAP) reduces the success of trabeculectomy.^16^ The present findings, showing that small palpebral fissures—clinically correlated with PAP—adversely affect postoperative outcomes, are therefore consistent with these earlier observations. To our knowledge, no prior studies have quantitatively defined small or narrow palpebral fissures as a preoperative risk factor for glaucoma surgery. Although one XEN-45 case series mentioned that a narrow palpebral fissure may have limited bleb expansion and contributed to postoperative scarring in an individual case,^17^ this observation was anecdotal and not supported by statistical analysis.

On the other hand, logistic regression of commonly recognised background factors such as preoperative IOP or glaucoma disease type for glaucoma surgery (**Supplemental Table 3**) revealed that none were individually significant risk factors. This likely reflects real-world clinical decision-making: surgeons recognise patients who may be difficult to treat and proactively select more effective procedures, such as AGV implantation or trabeculectomy, for challenging cases such as exfoliation glaucoma and other secondary glaucoma.

As shown in Table 2, the AGV group had the highest prevalence of diverse background risk factors, demonstrating how surgeons tailor their choice of procedure to maximise success. Nevertheless, the fact that small palpebral fissures remained an independent risk factor despite such adjustments suggests that their influence is particularly strong and clinically relevant.

An interesting finding is that patients with background factors that typically prompt surgeons to choose minimally invasive procedures actually exhibited poorer outcomes. For example, in patients with mental illness or hearing loss, less invasive procedures such as Preserflo or MIGS tended to be selected due to concerns about postoperative management. However, in this study, the µTLO success rate for patients with these background factors was substantially lower than our previous report^6, 7, 11^ and the overall success rate of µTLO in this study, which was approximately 65%. This suggests that choosing a less invasive procedure to reduce perioperative burden may inadvertently increase the risk of treatment failure and subsequent reoperation. Clinically, this highlights the important message that selecting a “gentler” procedure does not necessarily lead to “better outcomes.”

Additionally, MIGS is based on the principle of avoiding bleb formation. Despite this, the success rate of µTLO in patients with small palpebral fissures was low at 55%. This likely reflects the clinical scenario in which surgeons “attempt MIGS first” because filtration surgery is technically difficult and expected to have low success in patients with small palpebral fissures. These findings highlight the limitations of µTLO in this subgroup. Although logistic regression also identified hearing loss as significant, it is unlikely that hearing loss directly affects ocular physiology. A more plausible interpretation is that hearing loss reflects systemic frailty or reduced functional reserve, ^18^ and that the patient’s general condition and lifestyle may influence postoperative outcomes. The small number of hearing-loss cases limited the feasibility of conducting propensity score analyses.

It should be noted that the non-significant results for other background factors in this study do not imply that these factors lack clinical importance;^19^ rather, their effects may be mitigated by surgeons’ appropriate selection of surgical procedures. For example, large database studies have demonstrated that atopic dermatitis is a risk factor for postoperative complications in glaucoma surgery.^20^ The fact that small palpebral fissures alone showed a significant effect after adjusting for surgeons’ decision-making emphasises the strong clinical impact of this factor.

This study has several limitations. First, it was a single-centre study conducted in Japanese patients, and validation in other ethnic populations is needed. Second, multiple surgical procedures were analysed simultaneously, and larger sample sizes will be required to evaluate each procedure in greater detail. Third, the use of different visual field test devices limited the ability to quantitatively compare preoperative glaucoma severity.

In summary, this study revealed that many background factors in glaucoma surgery are often overlooked. Using both logistic regression and propensity score matching, we also demonstrated that small palpebral fissures are a strong risk factor for surgical failure.

Surgeons should recognise that patients with small palpebral fissures generally present with more challenging clinical profiles, which may reduce the success of both MIGS and filtration surgery, and should account for this in surgical decision-making. Ophthalmologists primarily involved in non-surgical glaucoma management should also be aware that chronic use of glaucoma medications, particularly prostaglandin F receptor agonists that may induce blepharitis and deepening of the upper eyelid sulcus,^21, 22^ can influence future surgical outcomes and should consider long-term implications when planning treatment.

## Data Availability

All data produced in the present study are available upon reasonable request to the authors.

## Notes

### Competing Interest Statement

The authors have declared no competing interest.

### Author Declarations

The study adhered to the tenets of the Declaration of Helsinki and was approved by the institutional review board of Kobe University Hospital (No. B250117).

