## Supplemental Table 1 for "Small palpebral fissure as a significant risk factor for glaucoma surgery failure"

Supplemental Table 1. Surgical success rates according to factors tot commonly considered risk Factors

|  | All | µTLO | Preserflo | Trab | AGV |
| --- | --- | --- | --- | --- | --- |
| Very elderly age (≥85 Years), n=19 | 16,3 (84%) | 1,1 (50%) | 9,2 (82%) | 4,0 (100%) | 2,0 (100%) |
| Avitreous, n=18 | 16,2 (89%) | 0,1 (0%) | 4,0 (100%) | 1,0 (100%) | 11,1 (92%) |
| Aphakia, n=2 | 1,1 (50%) | 0,0 (-) | 0,0,(-) | 0,0 (-) | 1,1 (50%) |
| Antithrombotic agents, n=59 | 51,8 (86%) | 4,3 (57%) | 36,5 (88%) | 3,0 (100%) | 8,0 (80%) |
| Difficulty with frequent visits, n=74 | 59,15 (80%) | 6,5 (55%) | 39,8 (83%) | 9,1 (90%) | 5,1 (83%) |
| Small palpebral fissure, n=128 | 93,35 (73%) | 17,14 (55%) | 57,15 (79%) | 14,4 (78%) | 5,2 (71%) |
| Corneal endothelial disorder, n=10 | 9,3 (75%) | 0,1 (0%) | 2,2 (50%) | 2,0 (100%) | 5,0 (100%) |
| Poor vision in fellow eye, n=67 | 57,10 (85%) | 7,4 (64%) | 35,4 (90%) | 8,2 (80%) | 7,0 (100%) |
| Dementia, n=5 | 4,1 (80%) | 0,0 (-) | 4,0 (100%) | 0,1 (0%) | 0,0 (-) |
| Hearing loss, n=12 | 8,4 (67%) | 1,2 (33%) | 6,2 (75%) | 1,0 (100%) | 0,0 (-) |
| Mental illness, n=19 | 15,4 (79%) | 4,2 (67%) | 9,1 (90%) | 2,1 (67%) | 0,0 (-) |
| Atopic dermatitis, n=22 | 14,8 (64%) | 3,3 (50%) | 8,3 (73%) | 2.0 (100%) | 1,2 (33%) |
| Pseudophacodonesis, n=4 | 4,0 (100%) | 1.0 (100%) | 1,0 (100%) | 2,0 (100%) | 0,0 (-) |
| Glaucoma eye drop allergy, n=107 | 83,24 (78%) | 15,12 (56%) | 51,8 (86%) | 14,3 (82%) | 3,1 (75%) |
| Asthma / Heart Disease, n=50 | 39,11 (78%) | 7,3 (70%) | 22,6 (79%) | 5,1 (83%) | 5,1 (83%) |

µTLO, Microhook ab interno trabeculotomy: Trab: Trabeculectomy: AGV, Ahmed Glaucoma Valve: The left shows the number of successful surgeries, the right shows the number of unsuccessful surgeries, and the number in parentheses shows the success rate.
