## Supplemental Table 2 for "Small palpebral fissure as a significant risk factor for glaucoma surgery failure"

Supplemental Table 2. Factors not commonly considered general glaucoma risk factors in the small vs. large palpebral fissure groups by propensity score matching analysis

|  | Small palpebral fissure  (n=111) | Large palpebral fissure  (n=111) | P value |
| --- | --- | --- | --- |
| Very elderly age (≥85 Years) | 6 | 7 | 1.00 |
| Aviterous | 3 | 7 | 0.33 |
| Aphakia | 0 | 0 | 1.00 |
| Antithrombotic agents | 23 | 18 | 0.49 |
| Difficulty with frequent visits | 22 | 30 | 0.27 |
| Corneal endothelial disorder | 5 | 2 | 0.45 |
| Poor vision in fellow eye | 30 | 19 | 0.11 |
| Dementia | 3 | 1 | 0.62 |
| Hearing loss | 4 | 4 | 1.00 |
| Mental illness | 8 | 4 | 0.37 |
| Atopic dermatitis | 8 | 8 | 1.00 |
| Pseudophacodonesis | 0 | 3 | 0.25 |
| Glaucoma eye drop allergy | 32 | 38 | 0.47 |
| Asthma / Heart Disease | 22 | 14 | 0.20 |
