## Supplemental Table 3 for "Small palpebral fissure as a significant risk factor for glaucoma surgery failure"

Supplemental Table 3. Logistic regression analysis of surgical success in relation to common glaucoma risk factors

|  | Univariate analysis | | |
| --- | --- | --- | --- |
|  | Odds ratio | 95%CI | P value |
| Right | 1.4 | 0.82-2.38 | 0.22 |
| Glaucoma disease type (Compared to POAG) |  |  |  |
| XFG | 1.44 | 0.74-2.83 | 0.29 |
| Others | 1.97 | 0.98-3.94 | 0.056 |
| Combined cataract surgery | 0.88 | 0.49-1.55 | 0.65 |
| Surgeon (Compared to A) |  |  |  |
| Surgeon B | 0.81 | 0.39-1.67 | 0.56 |
| Surgeon C | 1.00 | 0.44-2.28 | 0.10 |
| Surgeon D | 0.82 | 0.40-1.72 | 0.61 |
| Preoperative IOP | 1.03 | 1.01-1.06 | 0.01 |
| Preoperative GDS | 1.10 | 0.92-1.31 | 0.31 |
| Number of preoperative intraocular surgery | 1.10 | 0.86-1.42 | 0.43 |
| Axial length | 0.99 | 0.85-1.14 | 0.86 |

POAG, Primary Open Angle Glaucoma: XFG, Exfoliation glaucoma: IOP, Intraocular Pressure: GDS: Glaucoma Drug Score
